# Sample Size Analysis for Machine Learning Clinical Validation Studies

**DOI:** 10.1101/2021.10.26.21265541

**Authors:** Daniel M. Goldenholz, Haoqi Sun, Wolfgang Ganglberger, M. Brandon Westover

**Affiliations:** Beth Israel Deaconess Medical Center, Boston USA; Harvard Medical School, Boston USA; Massachusetts General Hospital, Boston USA

**Keywords:** statistics, machine learning, power calculation

## Abstract

**OBJECTIVE:** Before integrating new machine learning (ML) into clinical practice, algorithms must undergo validation. Validation studies require sample size estimates. Unlike hypothesis testing studies seeking a p-value, the goal of validating predictive models is obtaining estimates of model performance. Our aim was to provide a standardized, data distribution- and model-agnostic approach to sample size calculations for validation studies of predictive ML models.

**MATERIALS AND METHODS:** Sample Size Analysis for Machine Learning (SSAML) was tested in three previously published models: brain age to predict mortality (Cox Proportional Hazard), COVID hospitalization risk prediction (ordinal regression), and seizure risk forecasting (deep learning). The SSAML steps are: 1) Specify performance metrics for model discrimination and calibration. For discrimination, we use area under the receiver operating curve (AUC) for classification and Harrell’s C-statistic for survival models. For calibration, we employ calibration slope and calibration-in-the-large. 2) Specify the required precision and accuracy (≤0.5 normalized confidence interval width and ±5% accuracy). 3) Specify the required coverage probability (95%). 4) For increasing sample sizes, calculate the expected precision and bias that is achievable. 5) Choose the minimum sample size that meets all requirements.

**RESULTS:** Minimum sample sizes were obtained in each dataset using standardized criteria.

**DISCUSSION:** SSAML provides a formal expectation of precision and accuracy at a desired confidence level.

**CONCLUSION:** SSAML is open-source and agnostics to data type and ML model. It can be used for clinical validation studies of ML models.

## Background and Significance

Reports of opportunities for machine learning (ML) to improve clinical care are being published at an accelerating rate. Proof-of-concept studies with a clinical ML model are now common. However, before adopting such models into clinical practice, they should undergo *validation*. Clinical validation provides a confirmation that the proposed algorithm can generalize to situations or patients not previously encountered, and helps to clarify limitations of an algorithm[1].

Clinical validation studies are often expensive, time consuming, and may expose subjects to risk. Therefore, researchers need to determine the minimum number of samples/events/patients needed to verify an algorithm with a specified confidence. Most clinical investigators are familiar with sample size calculations designed to test a yes/no question, couched in terms the familiar machinery of significance testing, i.e., the number of subjects or events needed to provide a given level of power to reject a null hypothesis with a specified level of type I error. By contrast, the goal of a validation study for a predictive model is to estimate model *performance measures*. That is, the role of a clinical validation study is to measure model performance - accurately and precisely. Accuracy means low bias (small distance from “true” estimates). Precision means high certainty (small confidence intervals). Thus, the goal of a clinical validation is conceptually distinct from hypothesis testing[2].

In addition, because ML models for medical problems can be complex, and increasingly deal with data generated by processes not well described by conventional statistical models, sample size calculations are not always straightforward for predictive models developed using ML techniques.

Here we propose a general algorithmic approach for Sample Size cAlculation for ML clinical validation studies (SSAML). SSAML makes no assumptions about the specific ML technique being validated, or about the type of data involved. This approach builds on earlier work by Collins et al[2]. We provide open-source computer code that implements SSAML, so that other investigators in translational medicine can use the technique to estimate the sample size needed for their own ML model validation studies.

## MATERIALS AND METHODS

The data were obtained in de-identified format from three previously published studies on neurologic prediction models. The first was a study of the Brain Age Index (BAI)[3], which used sleep EEGs from participants in the Sleep Heart Health Study[4]. The BAI study estimated the “brain age” based on a transformed linear regression model of EEG which was then compared to the participant’s actual age. Using survival analysis in combination with BAI, it was found that life expectancy could be estimated[5]. The second study (COVA) evaluated the risk of hospitalization, ICU admission, and death from COVID-19, utilizing an ordinal regression model[6]. The third study from Seizure Tracker™ (ST) developed a seizure forecasting system based on de-identified electronic seizure diaries using an artificial neural network model (deep learning)[7]. The ST study was determined exempted by the Beth Israel Deaconess Medical Center Institutional Review Board, 2017D000488. The COVA and BAI studies were determined exempted by the Mass General Brigham Institutional Review Board, 2013P001024. Basic characteristics of each study are summarized in Table 1.

**TABLE 1:** Basic characteristics of the 3 datasets used. BAI = brain age index. COVA = COVID-19 risk study. ST = Seizure Tracker™. In all 3 datasets, number of events and number of patients are listed, but only in the COVA dataset we employed an event-based analysis, whereas in BAI and ST we used a patient-based analysis.

| Dataset | Machine learning | # Of patients / # of events | Disease model | Outcome | Repeated measures | Survival analysis | Event-based analysis |
| --- | --- | --- | --- | --- | --- | --- | --- |
| BAI(3, 5) | Transformed linear regression | 4070 patients<br>3359 events | Aging | Estimate of brain age (used to forecast life expectancy) | N | Y | N |
| COVA(6) | Ordinal regression | 2205 patients<br>1479 events | COVID-19 | Risk of hospitalization, critical illness, or death | N | N | Y |
| ST(7) | Deep learning | 1613 patients<br>98119 events | Epilepsy | Risk of seizure within 24 hours | Y | N | N |

Our technique, SSAML consist of 5 steps, illustrated in algorithmic detail in Figure 1:

**FIGURE 1:**
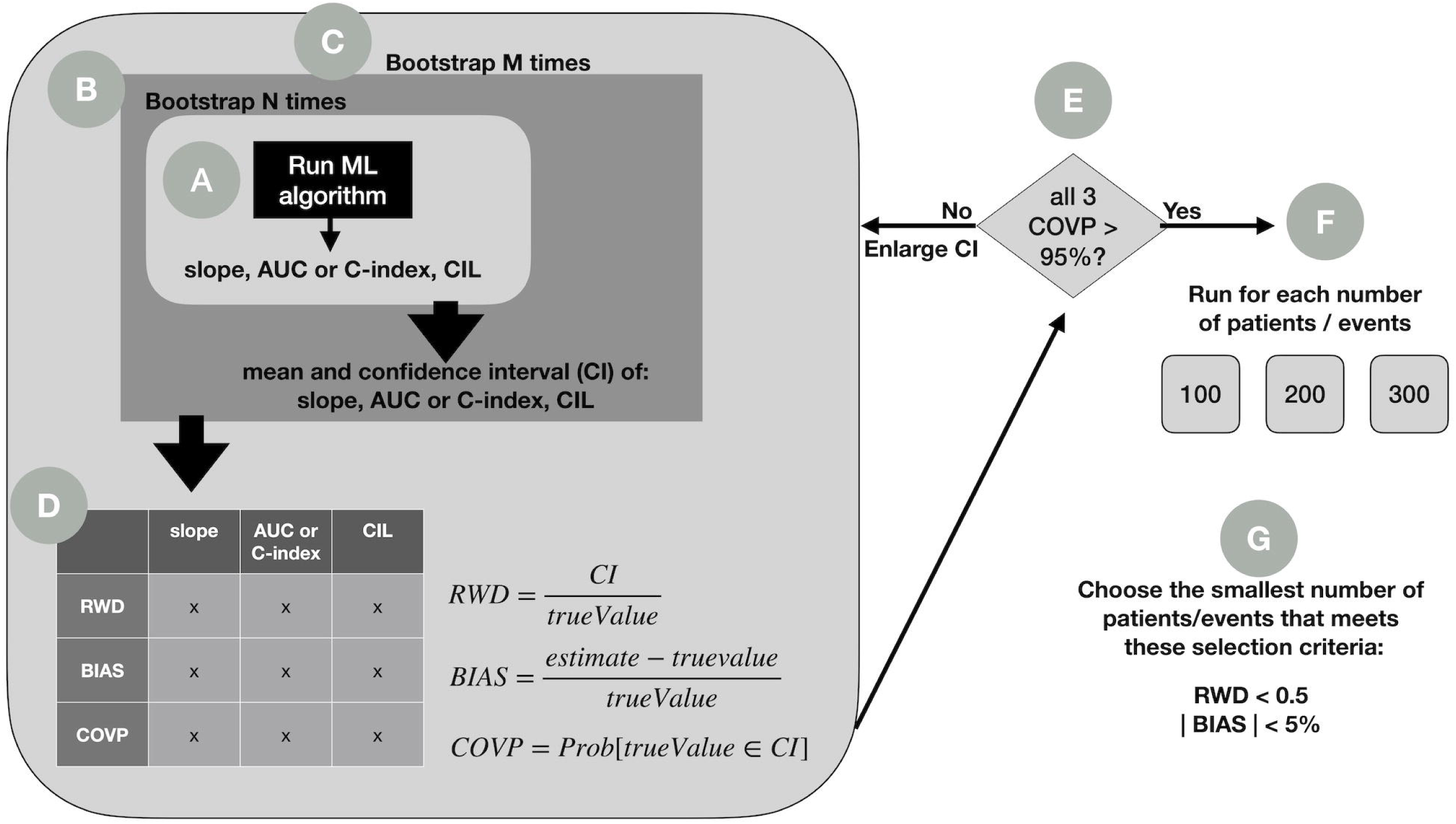
SSAML algorithm. A) The ML algorithm is run on a sample of data, and outcome metrics are computed. B) Step A is repeated N times from randomly chosen samples with replacement (i.e., bootstrapping), allowing computation of mean and confidence intervals from the outcome metrics. C) Step B is repeated M times (i.e., double bootstrapping) to obtain D) mean estimates of RWD and BIAS, as well as a point estimate for COVP. E) If COVP in any metric does not satisfy the 95% rule, the confidence interval (CI) is enlarged, and A through D are repeated. If all metrics meet the criteria, then F) this process can be repeated for additional number of patients (or events). In G) the smallest number of patients or events that satisfies all selection criteria is then chosen.

- **STEP 1)** Specify performance metrics, including measures of model discrimination (ability to distinguish cases from controls), and calibration (how well model risk predictions match observed case rates). For discrimination, we use the area under the receiver operating curve (AUC) for binary classification tasks (ST and COVA); we use the Harrell’s C-index (a generalization of AUC) for our survival model task (BAI). For calibration, we employ calibration slope and calibration-in-the-large (CIL)[8].
- **STEP 2)** Specify the required precision (relative width of confidence intervals, or RWD), and accuracy (percent bias or BIAS). We use cut-offs of ≤0.5 for precision and ±5% for accuracy.
- **STEP 3)** Specify the required confidence (probability that the CI includes the true value, i.e., “coverage probability” or COVP). We recommend 95%.
- **STEP 4)** For increasing sample sizes, calculate the expected precision and bias that is achievable, subject to the coverage probability requirement.
- **STEP 5)** Choose the minimum sample size that meets the requirements.

The ML approach to be validated is run on a bootstrapped sample of data, and the chosen discrimination and calibration metrics are computed; in our examples we used calibration slope, AUC or C-index, and CIL (Figure 1A). These calculations can be performed on real data or on simulated data that captures the investigator’s hypothesis about the data generating process. This calculation is repeated N times from randomly chosen samples with replacement (bootstrapping), allowing calculation of mean and confidence intervals from the performance metrics (Figure 1B). Bootstrapping is repeated M times (i.e., double bootstrapping[9], Figure 1C) to obtain mean estimates of RWD and BIAS, and for COVP (Figure 1D). If COVP<95% in any metric, the confidence interval (CI) is enlarged (Figure 1E), and the double bootstrapping procedure is repeated. If all metrics meet criteria, then (Figure 1F) this process is repeated with a larger number of patients (or events). The smallest number of patients or events that satisfies all selection criteria is then chosen (Figure 1G).

For each of our 3 illustrative datasets, we ran SSAML for a set of 4 possible sample sizes. The COVA dataset has a binary outcome measured once per patient, estimated using an ordinal regression. The BAI dataset has a numerical outcome measured once per patient with censorship (i.e., survival analysis), estimated using a Cox proportional hazard model. For BAI, we estimated the number of events needed for a validation study, rather than the number of patients needed. The ST dataset has a binary outcome which was measured in a timeseries and forecast repeatedly in each patient, estimated with deep learning. It is noted that the choice to use events rather than patients is at the discretion of the user – either way is essentially equivalent, but we opted to illustrate both.

In addition, we generated simulated 4 datasets to explicitly investigate the effect of different numbers of model input features (10 and 100), class imbalance ratios (1:1 and 1:10), and noise levels (input label were swapped at a rate of 10% and 20%) on the confidence intervals of the metrics from SSAML. These simulated datasets were fit with a logistic regression model, and SSAML was run on each.

Open-source code for SSAML and the simulations are available (https://github.com/GoldenholzLab/SSAML).

## RESULTS

The main features of each dataset are summarized in Table 1. Each dataset presents a unique challenge in terms of type of data, machine learning algorithm, and unique features. These examples were chosen to show the variety of applications SSAML can manage successfully.

The results from SSAML are summarized in Table 2. The results from the smallest confidence interval for each patient or event size that met the COVP≥95% condition is shown. Using this data table, it is possible to estimate a minimum sample size required for a clinical validation study that all meets the criteria established here (RWD<0.5, BIAS<5%, COVP>95%). These numbers are as follows: BAI, 1500 patients; COVA, 150 events; ST, 40 patients.

**TABLE 2:** Data tables from each dataset. Highlighted in bold are numbers that satisfy the requirements: RWD < 0.5, |BIAS| < 0.05, and COVP>0.95. The number of patients/events that satisfy the requirements for all categories are also highlighted in bold. Conf. int. = confidence interval. BAI = Brain Age Index (BAI) [3,5], COVA = COVID19 risk Assessment[6], ST = Seizure Tracker™[7]. Slope = calibration slope, AUC = area under the receiver operator curve, C-index = Harrell’s c-index, CIL = calibration-in-the-large, RWD = relative width of confidence interval, BIAS = bias in estimate compared with “true” value, COVP = probability of confidence interval covering “true” value. Note: for the confidence interval used for ST with 20 patients, the actual value was 0.9999, however due to rounding for three significant digits it is listed as 1.000.

| BAI |  | Number of participants |  |  |  |
| --- | --- | --- | --- | --- | --- |
| METRIC |  | 500 | 1000 | 1500 | 2000 |
| Conf.int. |  | 0.997 | 0.997 | 0.955 | 0.955 |
| RWD | slope | 1.602 | 0.848 | 0.444 | 0.382 |
| RWD | C-index | 0.157 | 0.110 | 0.061 | 0.052 |
| RWD | CIL | 0.196 | 0.134 | 0.073 | 0.063 |
| BIAS | slope | -0.053 | -0.024 | -0.008 | -0.008 |
| BIAS | C-index | -0.001 | -0.001 | -0.002 | -0.001 |
| BIAS | CIL | 0.005 | 0.002 | 0.002 | 0.001 |
| COVP | slope | 0.981 | 0.989 | 0.953 | 0.958 |
| COVP | C-index | 0.998 | 0.993 | 0.955 | 0.958 |
| COVP | CIL | 0.994 | 0.994 | 0.959 | 0.951 |

| COVA |  | Number of events |  |  |  |
| --- | --- | --- | --- | --- | --- |
| METRIC |  | 125 | 150 | 175 | 200 |
| Conf.int. |  | 0.955 | 0.955 | 0.955 | 0.955 |
| RWD | slope | 0.557 | 0.486 | 0.421 | 0.378 |
| RWD | AUC | 0.149 | 0.132 | 0.116 | 0.104 |
| RWD | CIL | 0.285 | 0.248 | 0.218 | 0.197 |
| BIAS | slope | -0.005 | 0.000 | 0.000 | 0.002 |
| BIAS | AUC | -0.001 | -0.001 | 0.000 | 0.001 |
| BIAS | CIL | -0.011 | -0.005 | -0.004 | -0.007 |
| COVP | slope | 0.966 | 0.964 | 0.956 | 0.973 |
| COVP | AUC | 0.968 | 0.979 | 0.972 | 0.974 |
| COVP | CIL | 0.977 | 0.971 | 0.966 | 0.977 |

| ST |  | Number of patients |  |  |  |
| --- | --- | --- | --- | --- | --- |
| METRIC |  | 20 | 40 | 60 | 80 |
| Conf.int. |  | 1.000 | 0.997 | 0.997 | 0.997 |
| RWD | slope | 1.001 | 0.324 | 0.187 | 0.131 |
| RWD | AUC | 0.378 | 0.205 | 0.168 | 0.146 |
| RWD | CIL | 0.370 | 0.165 | 0.129 | 0.107 |
| BIAS | slope | 0.076 | 0.022 | 0.010 | 0.007 |
| BIAS | AUC | 0.046 | 0.023 | 0.015 | 0.011 |
| BIAS | CIL | -0.026 | -0.010 | -0.008 | -0.005 |
| COVP | slope | 0.990 | 0.996 | 0.996 | 0.991 |
| COVP | AUC | 0.977 | 0.965 | 0.972 | 0.981 |
| COVP | CIL | 0.997 | 0.992 | 0.996 | 0.997 |

The 3 metrics for calibration and discrimination are plotted in Figure 2 for each of the 3 datasets. This plot illustrates the point that using increasingly larger sample sizes reduces the uncertainty (narrows the confidence intervals) for estimates of each of the metrics. In all 3 cases, the confidence intervals around slope narrow with increasing numbers of patients/events converging close to 1.0 as expected. The C-index converges to different final values depending on the dataset. BAI tended towards 0.6, COVA to just under 0.8, and ST to just under 0.9. These differences reflect the underlying discrimination capabilities of the specific models. The CIL values for BAI were considerably lower than ST and COVA. This, combined with slope, indicated better calibration for ST and COVA compared with BAI. Regardless of the differences in how accurate or precise these models may be, SSAML clarifies how many samples are needed for a validation study in each case.

**FIGURE 2:**
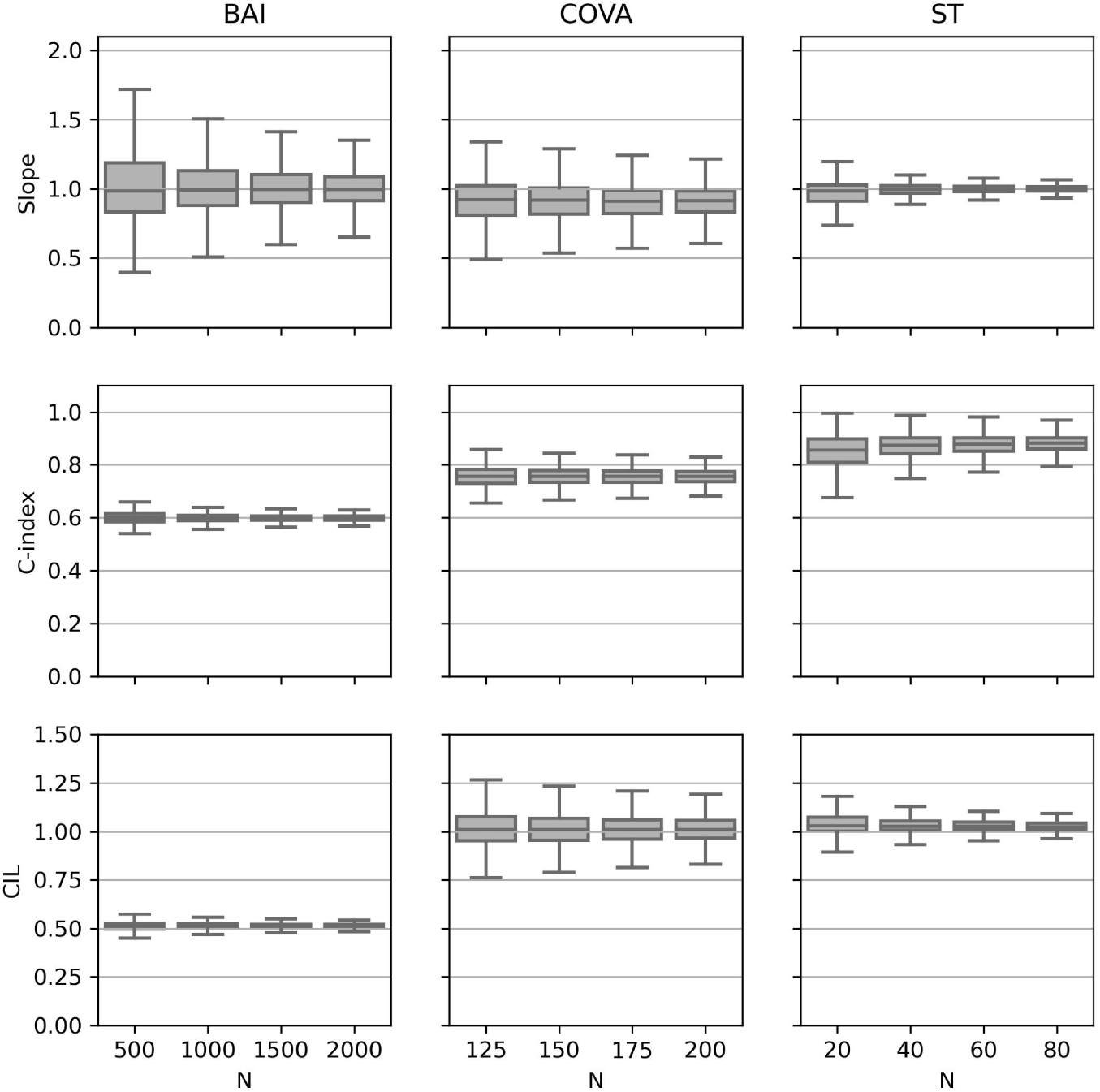
Narrowing confidence regions with increased number of patients/events. Shown here are 3 example datasets, one per column: Brain Age Index (BAI) [3,5], COVID19 risk Assessment (COVA) [6], and Seizure Tracker™ (ST) [7]. Each row indicates metrics for model performance: calibration slope (slope), area under the receiver operator curve or Harrell’s c-index (C-index), and calibration-in-the-large (CIL). In each subplot, as the number of patients or events (N) increases, the confidence interval narrows. When the desired performance level is reached, this represents the minimum powered study.

From the simulated datasets, several observations were made (Figure 3). First, the confidence intervals decreased with increasing N in all conditions. Next, the calibration slope was sensitive to the different conditions. Also, increased noise or input imbalance independently decreased the AUC. Additionally, CIL was not sensitive to different conditions tested. Finally, it is helpful to review slope, AUC and CIL to get a global sense of how well a given number of patients fits the model. Overall, the behavior of the simulated data helped to confirm intuitions about how SSAML would behave under various conditions.

**FIGURE 3:**
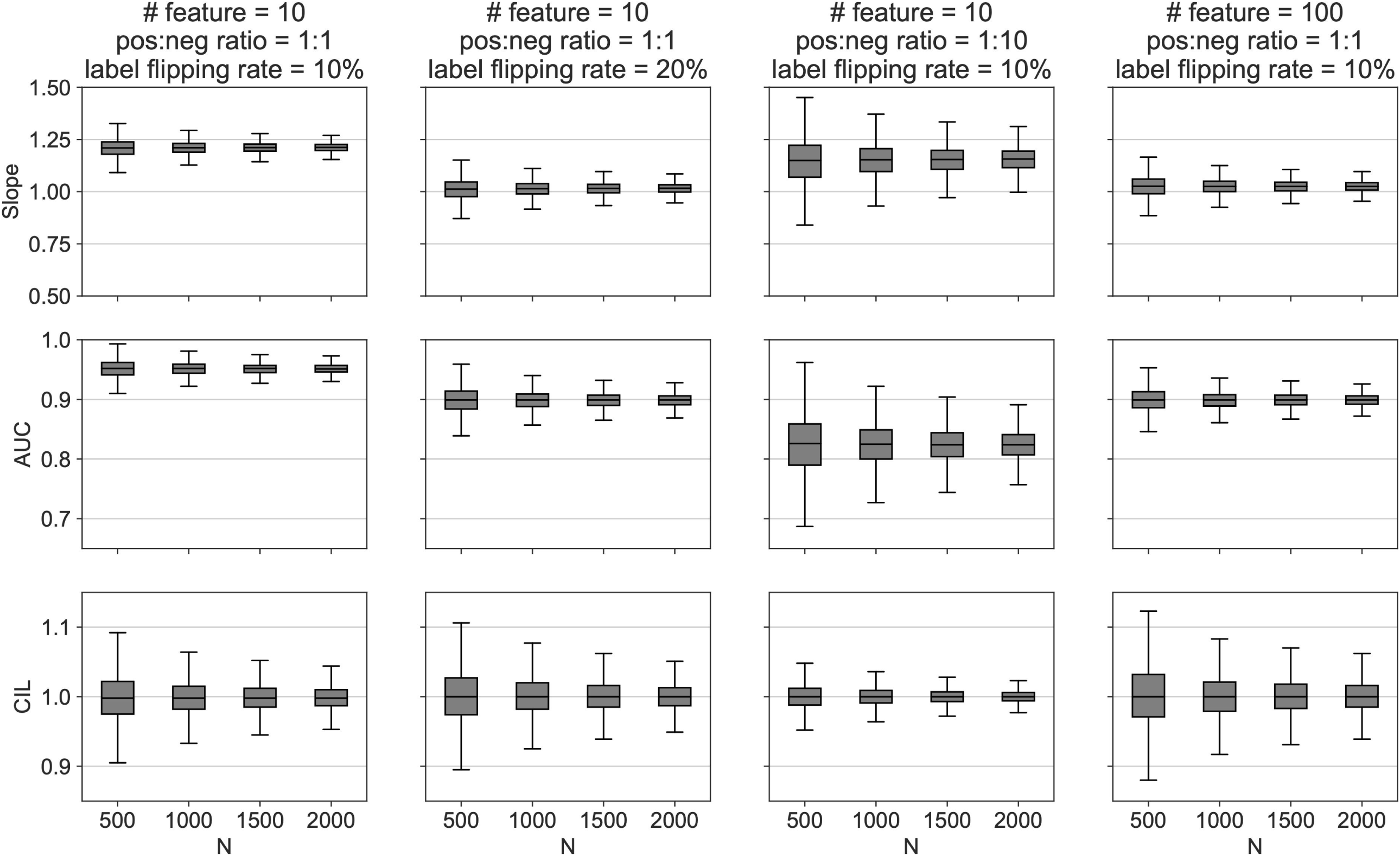
We generated 4 simulated datasets (in 4 columns) representing a binary classification problem that have different numbers of features (10 and 100), positive vs. negative class imbalance ratios (1:1 and 1:10), and noise levels (label flipping rate 10% and 20%). Each dataset was fit using a logistic regression. The fit data was then fed into the SSAML algorithm. The purpose is to explore the behavior of the confidence intervals of calibration slope (top row), AUC (middle row), and CIL (bottom row) under known conditions. Several simple observations can be made here: (1) the confidence intervals decreased with increasing N in all conditions; (2) the calibration slope was sensitive to different conditions; (3) increased noisy or class imbalance decreased the AUC; (4) CIL was not sensitive to different conditions tested; and (5) evaluating all 3 metrics (slope, AUC, and CIL) is important.

## DISCUSSION

As demonstrated here, SSAML provides algorithmic sample size calculations for validation of predictive models involving machine learning in clinical medicine. The methodology comes with several helpful advantages. First, it is agnostic to the specific machine learning techniques employed to develop predictive models. Second, it makes no assumptions about the underlying distribution of data. Third, it is available as an open-source tool. Fourth, it is flexible enough to manage single sample data (as in the case of COVA and BAI) or time series data (as in the case of ST). Finally, it provides precision and accuracy guarantees within pre-specified confidence ranges.

Each of the three datasets used highlights different features of SSAML (Table 1). For BAI and COVA, one sample was obtained per patient. For ST, there were multiple samples per patient (i.e., repeated measures). BAI was analyzed using survival statistics while COVA and ST were not. The number of events was computed for COVA, whereas for ST and BAI the number of patients was employed. Each of these three datasets were derived from different types of ML models. Finally, as seen in Figure 2, each of these models had different degrees of calibration and precision compared with the ground truth. In short, many different situations are well handled by SSAML.

The limitations of SSAML should be noted. Depending on the dataset, this method can be computationally expensive, and access to high performance computing may be beneficial. Moreover, if the sample of data is inadequate, or alternatively if one is unable to generate reasonable simulated data, SSAML cannot be expected to be accurate. If insufficient samples are included, or a large sample but not enough to fully capture the population distribution, both cases are expected to fail. An overly simplified example illustrates this point. Suppose an ML technique predicts migraine risk from eye movements. Using 1000 sighted participants as the input dataset for SSAML, the number of participants required for validation will be inaccurate if the validation study includes blind participants. As with any statistical model, the results of SSAML depend on the quality and representativeness of the data being used.

## CONCLUSION

SSAML is a general-purpose, model agnostic, open-source method for estimating sample sizes needed in clinical validation studies in clinical medicine involving a machine learning algorithm.

## Data Availability

All data produced in the present study are available upon reasonable request to the authors

https://github.com/GoldenholzLab/SSAML

## ACKNOWLEDGEMENTS

Portions of this research were conducted on the O2 High Performance Compute Cluster, supported by the Research Computing Group, at Harvard Medical School. See https://it.hms.harvard.edu/our-services/research-computing for more information.

